# Novel data preprocessing techniques in an expanded dataset improve machine learning model accuracy for a non-invasive blood glucose monitor

**DOI:** 10.1101/2023.07.24.23293113

**Authors:** Dominic Klyve, Kinara Pandya, Carl Ward, Barry Shelton

## Abstract

To determine the accuracy of a novel sensor designed to measure blood glucose (BG) non-invasively using Radio Frequency (RF) waves, we present results from a study that validates the stability of a machine learning model on an expanded dataset. In this study, we trained a Light Gradient-Boosting Machine (lightGBM) model to predict BG values using 3,311 observations from over 330 hours of data collected from 13 healthy participants, where an observation is defined as data collected from 13 sweeps from the novel Bio-RFID™ sensor paired with a single Dexcom G6® value as reference.

## 1. Introduction

Diabetes Mellitus is a condition characterized by high blood glucose (BG) that can result in severe long-term health consequences^1^; however, adherence with the practice of monitoring BG daily is poor^2^. Although modern continuous glucose monitors (CGMs) exist, these devices are not without limitations and come with the additional cost of regular replacement and discomfort of probe insertion. Adherence to monitoring is fundamental to effective treatment. Therefore, the development of a portable, non-invasive, and reliable point-of-care device for measuring BG is imperative.

In this report, we describe our efforts to employ novel data preprocessing techniques in our development of a machine learning model with improved accuracy for predicting BG. Data were collected with a new type of sensing device that rapidly scans through a wide band of RF frequencies and records values detected at each frequency over a period of time. We use readings of a Dexcom G6® as a proxy for BG, and predict values using a Light Gradient-Boosting Machine (lightGBM) model.

## 2. Methods

### 2.1 Data collection

Similar studies have been conducted in the past with 5 unique individuals of a similar demographic^3^. In the current study, data was collected from 13 healthy adults (4 female, 9 male) aged 24 – 61. Data were collected continuously over a 2 – 3 hour period with the patented Know Labs Bio-RFID sensor, using sweeps across the 500 MHz – 1500 MHz range at 0.1 MHz intervals, so each sweep collected data on 10,001 frequencies. Each sweep took approximately 22 seconds, including a one second pause between sweeps. Engineering details of this novel sensor are reported in a white paper^4^.

### 2.2 Data preprocessing

The dataset used in this analysis contained 3,311 observations from over 330 hours of data collected from 13 healthy participants across 110 two- to three-hour tests. In order to minimize noise in the data and to reduce the number of variables passed to the machine learning model, we maintained downsampling techniques in the temporal domain outlined in an earlier analysis on a smaller dataset^3^. In this analysis on an expanded dataset, we represented the frequency domain with more granularity by applying less aggressive feature reduction techniques of the RF spectrum. Here we reduced 10,001 spectral features to 167 by dividing each sweep into blocks of 60 frequency values and taking their mean, so that our data was in 6MHz intervals. In the previous analysis we downsampled to 40 spectral features^3^.

We also employed novel preprocessing techniques, including a focus on the differences of the sensor value between nearby frequencies, rather than the raw values returned by the sensor. More specifically, we replaced every frequency value with the difference between that frequency and the next (e.g., the difference given by 506MHz and 500MHz,etc.). We then used these differences as features in our machine learning model.

### 2.3 Model architecture and training

With the aim of predicting blood glucose based on RF waves we chose to explore Light Gradient-Boosting Machine (lightGBM) models, as we have previously found them to give good results with this type of data^5^. In a Python (version 3.10.11) environment, we conducted the training using the lightGBM package (version 3.3.5). When developing the lightGBM model, we employed a 60/20/20 train/validation/test split of the individual observations (readings from the CGM paired with averaged data from the sensing device over the same time period).

Hyperparameter tuning was conducted on the penalty terms and number of estimators, and the lowest Mean Absolute Relative Difference (MARD) achieved was taken for the final model, resulting at L1 = 0.4, L2 = 3, feature fraction = 0.5 and number of estimators = 2500. The model that yielded the best validation MARD was then used to perform a final evaluation on the test dataset to provide a ‘blind’ evaluation of model performance. Additionally, to assess model performance, we assessed the mean absolute error (MAE), and the percent of predictions that fell within 15%/20% of the reference Dexcom G6 glucose value.

## 3. Results

### 3.1: Comparing the training, validation and test datasets

We observed a MARD of 11.27% across the held-out test dataset. We also calculated a binary measure of success modeled after FDA limits for accuracy in new blood glucose monitors^6^. Each prediction is said to be “within threshold” if it is within 15% of the reference value. We found that 73.9% of values on the held-out test dataset were within threshold, as seen in Table 1.

**Table 1:** Results compared between training, validation, and test datasets. Error bars on the MARD and MAE give the 95% *t*-Confidence interval. Error bars on the ±15% and ±20% give the 95% *z*-Confidence interval for proportions.

|  | Observations | MARD (%) | MAE (mg/dl) | ±15% | ±20% |
| --- | --- | --- | --- | --- | --- |
| **Training** | 1986 (60%) | $0.03 \pm 0.001$ | $0.03 \pm 0.002$ | $100 \pm 0$ | $100 \pm 0$ |
| **Validation** | 662 (20%) | $11.17 \pm 0.79$ | $13.86 \pm 1.02$ | $73.72 \pm 3.35$ | $83.38 \pm 2.84$ |
| **Test** | 663 (20%) | $11.27 \pm 0.79$ | $13.76 \pm 1.01$ | $73.91 \pm 3.34$ | $82.05 \pm 2.92$ |

### 3.2: Comparing results across difference glycemic ranges

The model performs best in the normoglycemic range, with a MARD of 10.76%. The model performed well above chance in the hyperglycemic range with a MARD of 15.92%. Because our participant sample consisted of healthy adults, the dataset contained relatively few hyperglycemic reference values; these comprised only 8.3% of the test set. The training and validation sets had a similar proportion of normoglycemic to hyperglycemic values. There were insufficient observations in the hypoglycemic range in the test dataset. For a detailed comparison, see Table 2.

**Table 2:** Results broken down by glycemic status. Error bars on the MARD and MAE give the 95% *t*-Confidence interval. Error bars on the ±15% and ±20% give the 95% *z*-Confidence interval for proportions.

|  | Observations | MARD (%) | MAE (mg/dl) | ±15% | ±20% |
| --- | --- | --- | --- | --- | --- |
| **Hypoglycemic**<br>**(<70 mg/dl)** | 2 (<.3%) | n/a | n/a | n/a | n/a |
| **Normoglycemic**<br>**(70 – 180 mg/dl)** | 608 (91.4%) | 10.76 ± 0.79 | 12.00 ± 0.82 | 75.5 ± 3.4 | 83.6 ± 2.9 |
| **Hyperglycemic**<br>**(>180 mg/dl)** | 53 (8.3%) | 15.92 ± 2.98 | 33.43 ± 6.51 | 58.5 ± 13.3 | 67.9 ± 12.6 |

### 3.3 Clarke Error Grid analysis

We also performed a Clarke Error Grid analysis of our results. Developed in 1987 by Clarke et al.^7^, a Clarke Error Grid is a graphical representation used to assess the clinical accuracy of blood glucose measurement systems. The Clarke Error Grid analysis resulted in 82.4% of the blood glucose values falling into Zone A, 17.3% of the values in Zone B, 0% in Zone C, 0.3% in Zone D, and 0% in Zone E, as shown in Figure 1.

**Figure 1:**
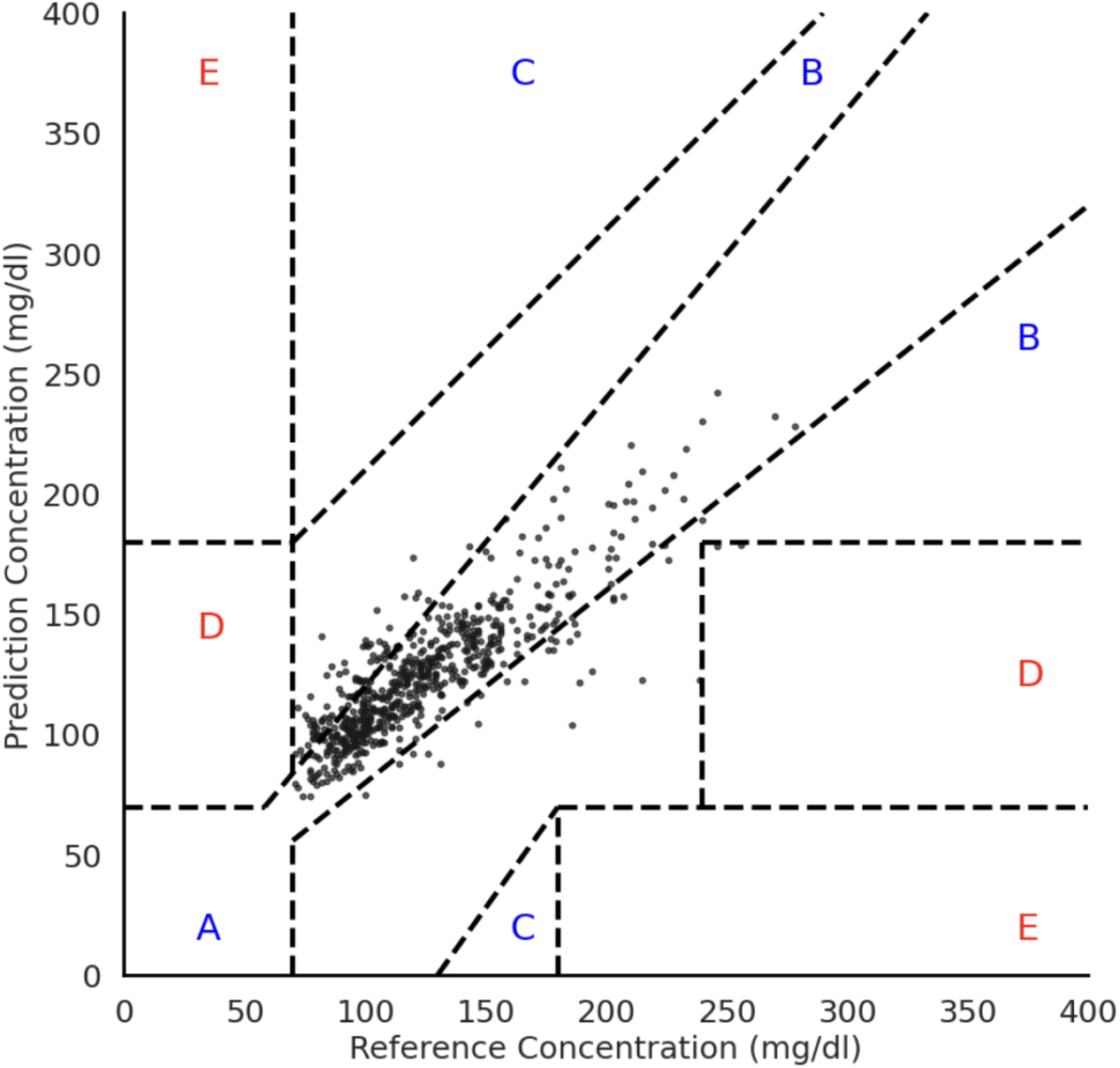
The Clarke Error Grid Analysis is depicted here to demonstrate which values in the test set fell into each error zone based on the glycemic range.

## 4. Discussion

This work demonstrates a test in which the patented Bio-RFID sensor was able to predict reference values of a gold standard CGM (Dexcom G6) continuously and non-invasively with a MARD of 11.27%. We describe the development of a method that employs novel aspects of data collection utilizing the Bio-RFID sensor and data preprocessing techniques to determine stability and reproducibility of a machine learning model trained on an expanded dataset. With a MARD of 11.27%, we have demonstrated stability of our model and an improvement in model accuracy when compared to previous test results^3^. At this stage, these developments support the use of such techniques as appropriate and feasible for larger scale testing of the validity of RF devices for BG management.

### 4.1 Limitations

Here we demonstrate the feasibility of a machine learning approach for non-invasive, RF-based BG detection systems. Before such technology is reliable for use in the real world, substantial implementation of our approach will be needed to support wider testing.

One limitation of this study is the requirement for a larger and more diverse participant population. All participants were healthy and did not have diabetes; indeed, 91.4% of the reference values were in the normoglycemic range. Future data collection and analysis must include expanded glucose ranges to understand our model’s robustness and ability to generalize to new individuals and across more glycemic ranges.

Another limitation is that our model was not designed to predict blood glucose directly, but rather to predict the values of a Dexcom G6 as a proxy for blood glucose, which itself has been independently assessed to have a MARD of 12.8% when compared to a Yellow Springs Instrument (YSI) device^8^, which is the clinical standard for direct measurement of BG. Moreover, there is a difference between the interstitial fluid measured by the reference device and the more complex muscle, blood, and interstitial space that the Bio-RFID sensor has access to. This suggests that our MARD of 11.27% may not be imprecision alone, but an artifact of the real differences between what is being measured.

In future work, it will be beneficial to explore the capability of the sensor to measure an entire cross-section of tissue, which could include arterial glucose, venous glucose, capillary glucose, interstitial glucose, and intracellular glucose. Moreover, further studies could eventually compare the Bio-RFID against a more precise blood glucose reference value, such as venous blood measured by a blood glucose reference analyzer.

## Data Availability

Data are produced in the present study are not available due to privacy and ethical concerns.

